# ReCo: a self-configuring and self-extending agentic framework for biomedical research

**DOI:** 10.64898/2026.07.14.26358025

**Authors:** Eleftherios Tzanis, Michail E. Klontzas

## Abstract

This study presents ReCo (Research Cosmos), a self-configuring and self-extending agentic research framework for the biomedical domain. ReCo is orchestrated by a large language model that interacts with native computing tools, bundled Model Context Protocol (MCP) servers, structured skills, persistent project memory, and a desktop interface. Its bundled MCP servers provide biomedical analysis capabilities while serving as implementation paradigms for integrating new computational and AI frameworks. Structured skills encode procedures for environment configuration and framework ingestion, enabling ReCo to inspect repositories, manuscripts, or local codebases; identify dependencies and execution patterns; create isolated runtime environments; design and implement MCP interfaces. Self-extension was evaluated using five heterogeneous systems: the Merlin computed tomography foundation model, MAISI-v2 medical image synthesis framework, asari liquid chromatography-mass spectrometry workflow, DosimeTron agentic radiation-dosimetry platform, and Orthanc DICOM server. ReCo successfully operationalized all five systems and completed predefined functional evaluations. Re-hosted DosimeTron outputs demonstrated near-perfect agreement with the reference pipeline across 651 organ observations (Pearson correlation and Lin concordance correlation coefficient, 0.99999; mean absolute percentage difference, 0.37%). Notably, ReCo configured Orthanc as a PACS-like coordination layer, integrated it with DosimeTron, Merlin, and TotalSegmentator, and orchestrated data retrieval, analysis, and return of valid DICOM RTSTRUCT, RTDOSE, and Structured Report. ReCo provides a unified environment for configuring, documenting, and operationalizing heterogeneous biomedical frameworks, reducing technical barriers to the adoption and integration of emerging computational and AI methods. The official open-source ReCo GitHub repository is available at: https://github.com/eltzanis/ReCo

## Introduction

Large language models (LLMs) have rapidly evolved from systems primarily associated with human-language generation into reasoning engines for agentic systems. In this setting, the model can select tools, generate and execute code, inspect files, query external resources, and use the results of these actions to continue a task. Early agentic approaches such as ReAct formalized the interconnection of reasoning and action, allowing language models to interact with external environments rather than only respond based on internal model knowledge [1]. Tool-use frameworks further demonstrated that language models can call external functions and application programming interfaces to extend their capabilities beyond text generation [2]. This transition is foundational for the development of systems that can automate complex, repetitive, and time-consuming computational processes. Recent medical-agent studies illustrate this shift from isolated language-model outputs toward systems that can participate in multi-step clinical workflows [3–5].

At the same time, AI-assisted software development has changed the pace at which code, analysis pipelines, and research frameworks can be produced. Coding agents can help implement software components, generate wrappers, debug errors, write documentation, and adapt code to new data or infrastructure. This acceleration creates an opportunity for research, but also a practical challenge. The number of newly released models and frameworks is increasing, while adoption often requires manual installation, dependency resolution, environment configuration, and framework-specific programming knowledge. As a result, even technically valuable tools may remain difficult to use, and researchers with limited programming experience can be excluded from advances that depend on rapidly changing software ecosystems. A major unmet need is therefore not only to develop more tools, but to develop systems that can help researchers discover, configure, understand, and operationalize those tools in a reliable and inspectable way, reducing the technical overhead required to deploy advanced computational methods and enabling researchers to focus on scientific discovery.

Agentic systems have used several approaches to expose computational capabilities to LLMs. In many early implementations, capabilities were provided as individual tools, often defined as Python functions or application-specific routines with structured inputs and outputs. This approach can be effective, but it couples each tool to a particular agent implementation and requires repeated custom integration. The Model Context Protocol (MCP) addresses this fragmentation by defining an open protocol for connecting AI systems to external data sources and tools through MCP servers and clients [6]. Because MCP provides a common interface, tools can be served independently of a specific agent framework and can be reused by different MCP-compatible clients. A complementary development is the use of skills, which represent structured instruction packages that teach an agent how to perform a recurring multi-step procedure. In contrast to a tool, which executes a specific operation, a skill provides procedural knowledge, such as how to inspect a codebase, follow a checklist, operate a framework, or generate outputs according to a defined workflow. Skills provide a mechanism for encoding operating procedures, while MCP servers provide executable capabilities.

In this study we present ReCo (Research Cosmos), a prototype self-configuring and self-extending agentic system for biomedical research. ReCo was designed as a research platform orchestrated by an LLM-based agent that combines native computing tools, bundled MCP servers, structured skills, project memory, and a desktop user interface. The bundled MCP servers provide immediate access to established biomedical research capabilities [7, 8], while also serving as implementation paradigms for the creation of new servers. Skills encode procedures for first-install configuration and ingestion of new frameworks, allowing the agent to inspect external repositories or local codebases, infer dependencies, propose tool surfaces, create isolated runtime environments, generate MCP wrappers, document the new capabilities, and register them for future use. The system was also designed to retain project-specific context through dynamic memory and session traces. In this work, we evaluated whether this architecture could extend ReCo beyond its original bundled toolkit by adding heterogeneous research frameworks, re-hosting an existing agentic system, connecting to imaging infrastructure, and orchestrating biomedical AI workflows through the generated interfaces.

## Materials and Methods

### Agentic System Architecture

ReCo was developed as an agentic research system built on the Claude Agent SDK [9], with a configurable large language model serving as the reasoning engine and a set of native tools enabling interaction with the local computing environment, including shell execution, file reading, file writing, targeted file editing, filesystem search, text search, web fetching, and web search. The system bundles a curated set of Model Context Protocol (MCP) [6] servers derived from previously developed research tools [7, 8]. These pre-installed servers provide immediate capabilities for exploratory data analysis (EDA), feature-importance analysis and feature selection, radiomic feature extraction, tabular classification and regression model development, medical image segmentation through TotalSegmentator [10] and nnU-Net [11], two-dimensional and three-dimensional medical image classification, model-performance comparison, and performance-decline monitoring and fine-tuning workflows. The central design goal was, however, not only to provide a fixed collection of predefined research tools, but to make the system capable of self-configuration and self-extension (Figure 1). In this design, the bundled MCP servers have a second function as reference implementations that guide the ingestion of new frameworks with different runtime patterns. ReCo uses structured skills to encode reproducible operating procedures for the agent. During first-install configuration, the setup skill guides the agent through detection of Conda, installation of Miniconda when required, creation of five isolated Conda [12] environments grouped by dependency compatibility, installation and verification of the required Python packages, and registration of the resulting interpreter paths for the bundled MCP servers. For extension beyond the bundled toolset, the user provides the agent with a Git repository or local source folder and requests in natural language that the framework be added to ReCo. Guided by the MCP-installation skill, the agent obtains the source following user confirmation, inspects dependency files and entry points, determines the required Python version, and proposes a tool surface for user approval. The agent then studies the bundled MCP servers as implementation paradigms, including synchronous analysis tools such as EDA, command-line wrappers such as TotalSegmentator, batch-processing tools, and long-running background-job patterns such as nnU-Net. These examples provide templates for JSON-schema tool definitions, stdout discipline required by MCP stdio communication, background execution for training or other long-running tasks, polling tools, model-cache handling, and documentation structure. After the user confirms the plan, the agent creates a dedicated Conda environment, installs dependencies, generates a new MCP wrapper and manifest, writes an accompanying helper module and documentation file, and verifies the server through an MCP handshake that sends ‘initializ’ and ‘tools/list’ requests before registration. The newly installed server is then added to the ReCo server catalog but left disabled by default, so that the user explicitly controls when it becomes available to future chats. ReCo also includes project-level memory, implemented as a persistent Markdown file injected into the agent context for each active project, allowing durable project-specific information to be retained across sessions. Agent behavior is further shaped by a ReCo-specific system prompt that defines the system identity, tool-use preferences, and project-memory behavior. The reasoning backend is configurable, with support for hosted and local model providers, including Anthropic [13], DeepSeek [14], local Ollama, and Ollama Cloud [15].

**Figure 1.**
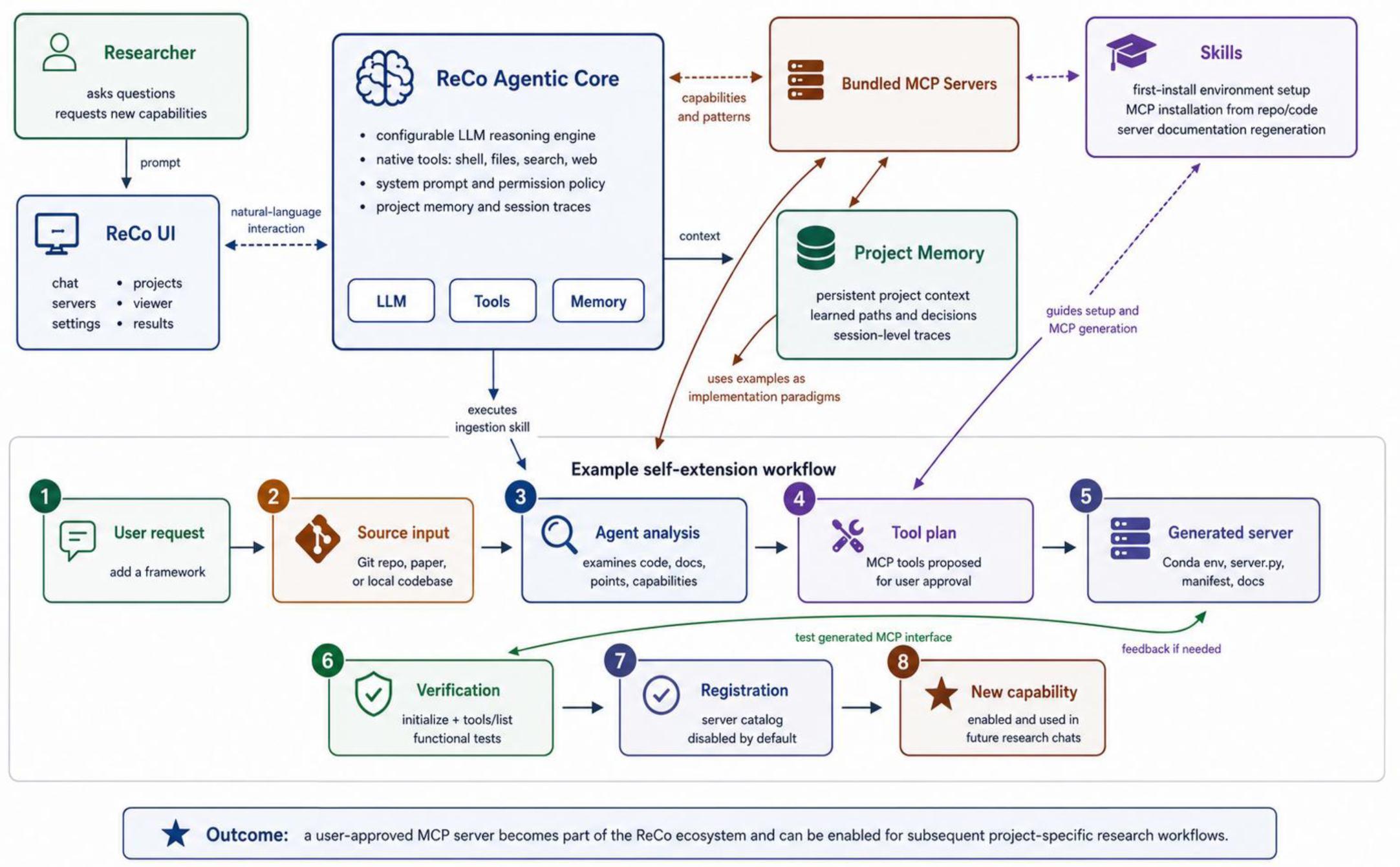
ReCo combines a configurable agentic core, bundled MCP servers, structured skills, project memory, and a desktop user interface. When a user requests a new capability from a repository, paper, or local codebase, ReCo analyzes the source, proposes an MCP tool surface, creates the runtime environment and server files under user supervision, verifies the generated MCP interface, and registers the new server for future project-specific workflows.

### Desktop Application and User Interface

The ReCo agentic system is exposed through a desktop application developed with Electron, Vite, React, and TypeScript [16, 17]. The application separates the main process, preload bridge, shared inter-process communication types, and renderer interface, allowing the agent runtime, MCP server catalog, settings, project state, session history, and image-viewer events to be managed by the main process while being presented through a graphical user interface (Figure 2). The packaged application includes the bundled Python MCP servers, server manifests, skills, user guide, and application assets as app resources, with MCP servers and skills unpacked to filesystem paths so that Python interpreters and subprocesses can access them at runtime. The interface is organized into task-specific panels. The Chat panel provides the natural-language interaction surface, streams agent responses, displays tool calls and tool outputs, and presents permission prompts when user approval is required. The Servers panel lists bundled and user-installed MCP servers by category, allows users to enable or disable servers, configure the Python interpreter associated with each server, inspect model-cache usage, open server documentation, install new servers from repositories or local folders, and remove user-installed servers. The Skills panel displays bundled and user-installed skills and allows them to be enabled or disabled. The Projects panel allows users to create named research workspaces, select the active project, edit the project memory file, and retain project-specific context across conversations. The Previous Chats panel lists stored sessions, supports filtering by project, and allows prior conversations to be reopened, renamed, exported, or deleted. These stored sessions also serve as behavioral traces of agent operation, since the chat system records user prompts, agent responses, tool invocations, and detailed tool inputs and outputs, allowing prior workflows and agent decisions to be reviewed after completion. The Settings panel provides configuration of the reasoning backend, model selection, native tool availability, and permission policy. The Viewer panel integrates a medical image viewer for anatomical images and segmentation masks, enabling visual inspection, overlay control, mask editing, and saving of corrected masks after agent-generated segmentation workflows. Together, these components expose the agentic system through a research interface in which users can configure the reasoning model, manage available tools, organize projects, supervise tool execution, inspect generated imaging outputs, and extend the system with additional MCP servers.

**Figure 2.**
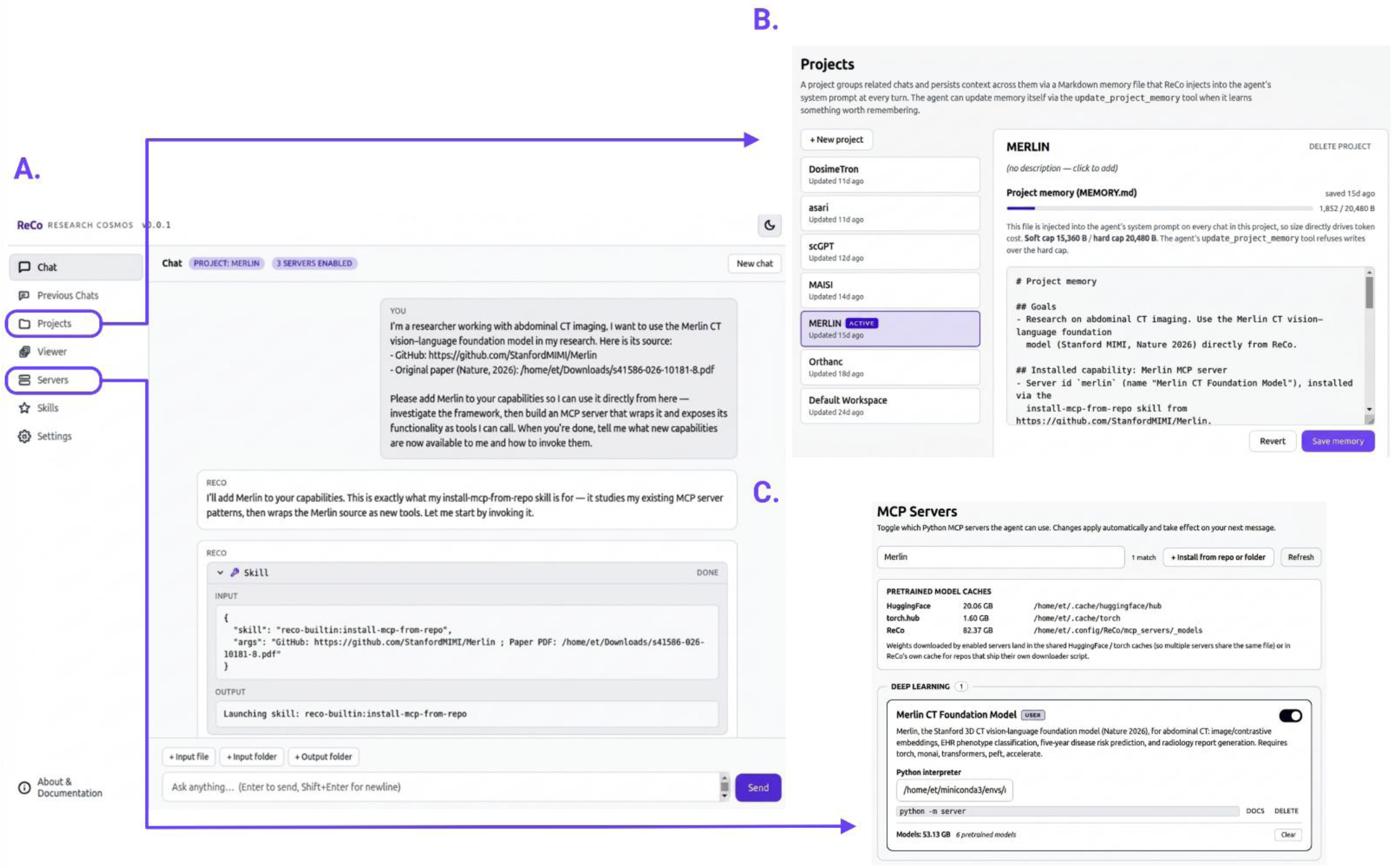
ReCo desktop application and graphical user interface. (A) Conversational onboarding of a new research framework, showing the progression from a natural-language capability request. (B) Project-management interface supporting persistent, project-specific context through an editable memory file that is retained across related chat sessions. (C) MCP server catalog, through which installed capabilities can be inspected, documented, activated, or disabled.

### Evaluation of Self-Extension Capability

The evaluation was designed to test whether ReCo could extend its own capability set by ingesting previously unseen research code/software and exposing it as usable MCP servers. The evaluation focused on capability acquisition and faithful encapsulation rather than re-benchmarking the underlying scientific frameworks. Five case studies were selected to cover distinct biomedical software and research-workflow archetypes: Merlin, a CT foundation model [18]; MAISI-v2 [19], a medical image synthesis framework; asari [20], a deterministic command-line workflow for liquid chromatography-mass spectrometry (LC-MS) metabolomics data processing; DosimeTron [21], a local agentic framework for personalized Monte Carlo (MC) radiation dosimetry in PET/CT; and Orthanc [22], a DICOM server used to evaluate infrastructure setup and real-world orchestration of AI workflows. For all validation experiments, Claude Opus 4.8 was selected as the core reasoning model of ReCo. For each case, the user supplied ReCo with a repository link, local source folder, and/or associated manuscript, and requested that the framework be added to the ReCo ecosystem. The prompts intentionally did not define the final MCP tool surface. This design allowed the evaluation to examine whether the agent could infer the operational structure of each framework, identify its dependencies, and construct an appropriate interface using the MCP-installation skill and the bundled MCP servers as implementation references.

For each framework-ingestion task, ReCo was expected to (i) inspect the source code, documentation, dependency files, and associated manuscript where available; (ii) determine an appropriate Python environment; (iii) create an isolated Conda environment; (iv) install the required dependencies; (v) design a tool surface; (vi) generate the MCP server implementation, manifest, helper files, and documentation; and (vii) register the resulting server in the application catalog after verification. The verification procedure included an MCP initialization request and ‘tools/list’ call when technically possible, followed by functional testing through new user prompts. The evaluation included frameworks with different integration patterns. Merlin required wrapping model-level Python functionality for CT image embedding, zero-shot finding classification, phenotype classification, cross-modal CT-report retrieval, five-year disease-risk prediction, and radiology-report generation. MAISI-v2 required handling pretrained generative models, model-cache organization, image-generation workflows, and GPU-memory constraints. asari required recognition and encapsulation of a command-line metabolomics workflow. DosimeTron tested whether ReCo could re-host an already agentic, multi-server dosimetry framework from a local codebase while preserving its original orchestration logic. Orthanc tested whether ReCo could configure external infrastructure, build a new MCP client for a DICOM server, and then use that infrastructure to orchestrate clinically realistic AI workflows involving DosimeTron, Merlin, and segmentation models through a PACS-like interface.

Evaluation prompts and expected capability targets were defined for each case before assessment, using the corresponding source paper, and project’s codebase documentation. The validation prompts assessed whether the generated MCP servers could execute representative framework functions, whether outputs were produced in the expected formats, and whether the agent respected framework boundaries when asked to perform tasks outside the supported scope. Negative-control prompts were included where appropriate to assess whether ReCo declined unsupported requests or overextended the framework. The resulting sessions were retained as chat transcripts containing user prompts, agent responses, tool calls, tool inputs, and tool outputs. Saved output artifacts were then inspected directly, and quantitative values were independently recomputed. This audit strategy was used to ground the evaluation in observable artifacts rather than narrative summaries alone. Results are reported separately for each framework, distinguishing functional encapsulation, quantitative reproducibility, qualitative output inspection, infrastructure setup, and orchestration performance according to the type of framework being evaluated.

### Datasets and Hardware

Each ingested framework was tested on data appropriate to its modality. For Merlin, test cases were drawn from the publicly released Stanford AIMI Merlin abdominal-CT dataset [18]: a main set of five studies (AC423ccbe, AC4240ff5, AC424421b, AC423e6ba, AC423f4f0) and a further five studies carrying five-year outcome labels (AC423ccbe, AC4216c26, AC4213e7c, AC42148eb, AC423bfcf). Zero-shot finding and five-year disease labels were taken from the dataset’s released label files. As an out-of-modality negative control, one abdominal MR volume was drawn from the publicly available TotalSegmentator-MRI dataset [23]. For asari, we used the authors’ publicly distributed example LC-MS data (https://github.com/shuzhao-li-lab/data/tree/main/data). For DosimeTron, six PSMA PET/CT studies (three ¹⁸F-PSMA and three ⁶⁸Ga-PSMA acquisitions) from distinct patients were selected from the whole-body PSMA-PET/CT dataset of Jeblick et al. [24]. The ground truth organ/tissue radiation doses for the dosimetric comparison was DosimeTron’s MC results on the same six cases. Ethics committee approval was not required for this study because all datasets used were publicly available, open-source datasets containing de-identified data.

ReCo was developed and tested on a single workstation equipped with an AMD Ryzen 7 5800 CPU, 64 GB of RAM, and an NVIDIA RTX 2080 Ti GPU (11 GB memory).

## Results

### Self-Extension Across Heterogeneous Research Frameworks

ReCo was evaluated on five framework-ingestion tasks (Table 1) that differed in software structure, dependency profile, execution pattern, and scientific domain. Across Merlin, Maisi-v2, asari and DosimeTron frameworks, ReCo generated usable MCP interfaces for the source frameworks and completed the predefined functional tests through the generated framework-specific servers (Table 2). Orthanc was evaluated as an infrastructure and orchestration case: ReCo configured a DICOM server, generated an MCP client for it, and used the resulting PACS-like environment to execute complex workflows involving DosimeTron, Merlin, and TotalSegmentator. The evaluated cases therefore tested self-extension across model-level Python APIs, generative imaging pipelines, deterministic command-line scientific software, a local multi-server agentic dosimetry framework, and external clinical-imaging infrastructure. For each included case, ReCo identified a runnable dependency configuration, created the required Conda environment or external service configuration, and executed the validation prompts through the generated interface. Runtime or configuration issues did not prevent workflow completion; when they occurred, ReCo diagnosed and corrected them.

**Table 1.** Framework-ingestion cases used to evaluate ReCo self-extension.

| **Case** | **Framework type** | **ReCo-generated interface** | **Evaluation focus** |
| --- | --- | --- | --- |
| **Merlin** | CT vision-language foundation model | Single MCP server with tools for CT image embedding, cross-modal CT-text scoring/retrieval, phenotype classification, five-year disease prediction, report generation, and job monitoring | Functional exposure of model-level CT capabilities |
| **Maisi-v2** | 3D medical image synthesis framework | Single MCP server with tools for model download, image generation, paired mask-image generation, mask-conditioned generation, and job monitoring | Qualitative and format-level validation of generated CT and brain MR outputs |
| **asari** | Deterministic LC-MS metabolomics command-line workflow | Single MCP server with processing, analysis, annotation, information, and job-monitoring tools | Reproduction of core command-line processing and artifact-level recomputation |
| **DosimeTron** | Local multi-server agentic dosimetry framework | Four MCP servers preserving the original DICOM, preprocessing, GATE, and dosimetry separation | Re-hosting fidelity for PET/CT dosimetry workflows |
| **Orthanc** | DICOM server and PACS-like infrastructure | Single MCP client server for Orthanc REST operations and background jobs | Infrastructure setup and real-world orchestration with DosimeTron, Merlin, and segmentation tools |

**Table 2.** Validation outputs generated after ReCo ingestion.

| **Case** | **Generated validation outputs** | **Verification approach** |
| --- | --- | --- |
| **Merlin** | CT embeddings, CT-text finding scores, phenotype predictions, disease-risk scores, CT-report retrieval rankings, and generated radiology-report text | Tool-output inspection, expected-positive ranking checks, report-structure review, and CT-domain negative control |
| **Maisi-v2** | CT NIfTI volumes, paired CT image-label outputs, mask-conditioned CT outputs, tumor-containing CT image-label outputs, geometry-controlled CT volumes, and brain MR volumes | File-format, geometry, intensity-range, label-alignment, and visual output review |
| **asari** | Feature tables, quality-control summaries, empirical-compound annotations, mass-track links, extracted-ion chromatogram values, and reproducibility summaries | Direct artifact inspection and independent recomputation from generated output tables |
| **DosimeTron** | Organ-level dose reports, segmentation-derived organ statistics, Monte Carlo outputs, and dosimetry summaries | Quantitative comparison with original DosimeTron agentic outputs across matched cases |
| **Orthanc** | Uploaded and retrieved DICOM studies, RTSTRUCT objects, RTDOSE objects, DICOM Structured Reports, secondary-capture objects, and encapsulated documents | Orthanc inventory checks, DICOM validity checks, downstream workflow execution, and DosimeTron arm comparison with direct file-based runs |

### Foundation Model Framework Ingestion and Functional Testing

For Merlin, ReCo inferred and exposed the evaluated CT foundation-model capabilities as MCP tools without being given an explicit tool specification. The generated interface supported CT image embedding, cross-modal CT-text scoring and retrieval, phenotype classification, five-year disease-risk prediction, radiology-report generation, and job-status monitoring. The predefined tests covered six evaluated Merlin capabilities: CT image embedding, zero-shot finding classification using CT-text similarity, phenotype classification, cross-modal CT-report retrieval, five-year disease-risk prediction, and report generation. These requests were executed through the generated Merlin MCP server on abdominal CT inputs from the Merlin evaluation data. ReCo produced the expected output types for the tested tasks, including CT embeddings, CT-text similarity scores, phenotype-classification outputs, disease-risk scores, candidate-report rankings, and generated report text. In the zero-shot finding-classification task, the expected positive findings were ranked first for the tested examples; in the cross-modal retrieval task, the true report was ranked first among the candidate reports. The five-year disease-risk task produced higher mean scores for positive than negative disease instances in the small validation set. These task-level findings were used to assess whether ReCo had correctly exposed the Merlin functions, not to re-benchmark Merlin. A negative-control prompt using an abdominal MRI input was rejected as outside the CT-specific operating domain. Report generation encountered GPU memory exhaustion during testing; ReCo responded by re-running the job on CPU, which allowed the workflow to complete.

### Synthetic Medical Image Generation Framework Ingestion and Functional Testing

For MAISI-v2, ReCo created an MCP server that exposed model acquisition and asynchronous generation workflows for CT and brain MR synthesis. The generated tool surface included paired image-mask generation, image-only generation, mask-conditioned generation, model download, and job monitoring. The evaluation tested image-only CT generation, paired CT label-map and image generation, CT generation from a supplied mask, hepatic tumor insertion, output geometry control through spacing and field-of-view settings, and brain MR generation with different image weightings (T1- and T2-weighted). All requests were executed through the generated MAISI-v2 MCP server, producing NIfTI image and label-map files. The generated CT and MR volumes were inspected for expected dimensions, voxel spacing, datatype, intensity range, label-image alignment, and visually identifiable requested structures where applicable. During evaluation, GPU memory limitations required user-approved tiling changes to the inference configuration; these changes modified memory handling rather than the requested output geometry or model task. A negative-control prompt requesting mask-conditioned brain MR generation was declined because the available ControlNet pathway was CT-specific. The MAISI-v2 results therefore support qualitative and workflow-level encapsulation of the released generative framework, but do not constitute a quantitative image-quality benchmark.

### LC-MS Metabolomics Framework Ingestion and Functional Testing

For asari, ReCo recognized the framework as a command-line LC-MS metabolomics workflow and generated an MCP interface around the relevant processing, analysis, annotation, information, and job-monitoring operations. The evaluation used the SZ22 example dataset and tested full dataset processing, single-file analysis, quality-control summary extraction, empirical compound annotation, feature trackability, and reproducibility across replicate and split-processing workflows. ReCo executed the core processing and analysis requests through the generated asari MCP server and produced feature tables, quality-control summaries, empirical-compound annotations, mass-track links, extracted-ion chromatogram values, and reproducibility summaries (Figure 3). Generated artifacts were inspected directly, and key quantities were recomputed from the output tables, including feature counts, quality-control fractions, empirical-compound counts and selected extracted-ion chromatogram values. The resulting values matched the saved artifacts, supporting faithful execution of the core asari workflow through ReCo.

**Figure 3.**
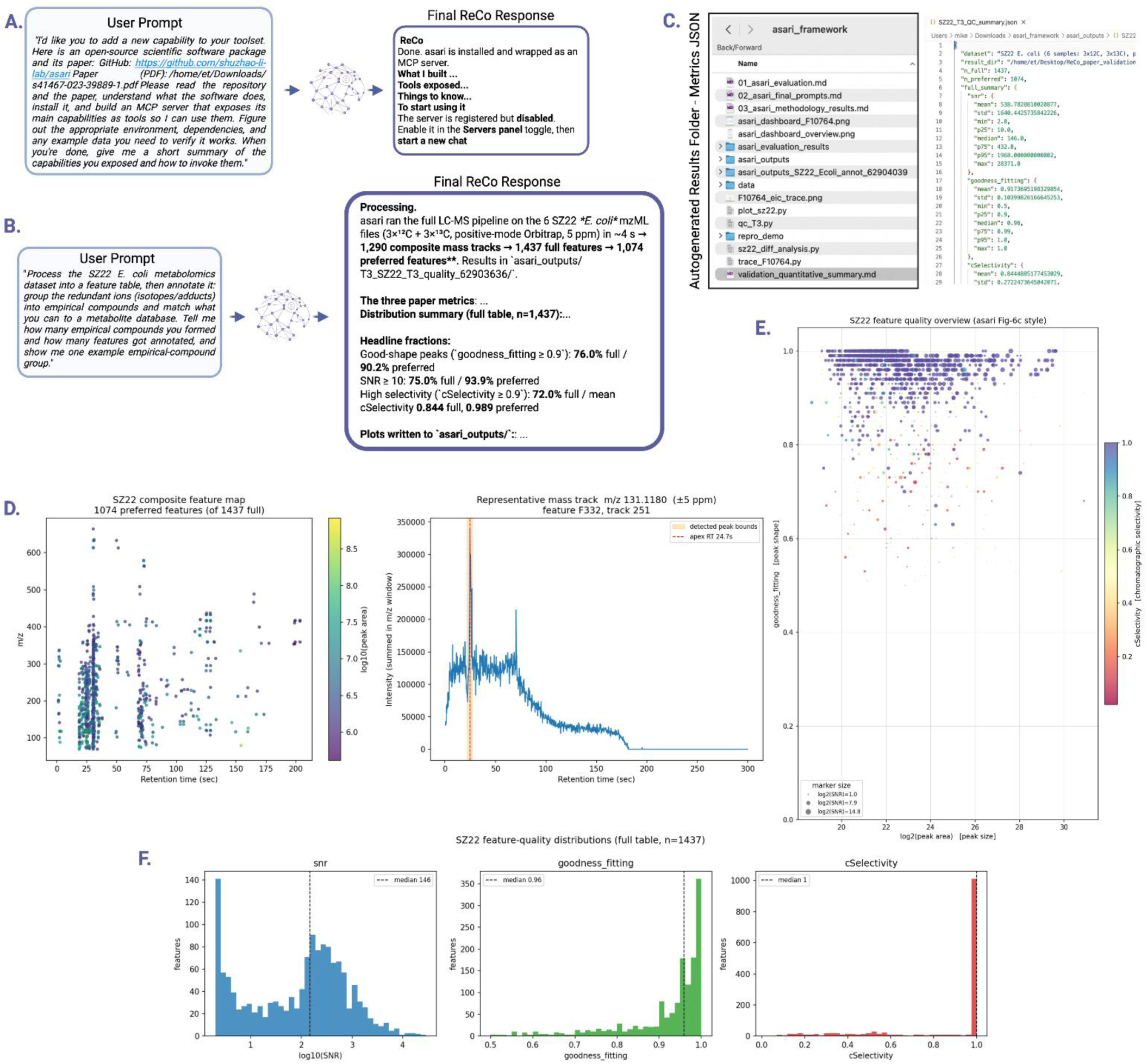
Validation of the automatically generated asari MCP server through representative metabolomics workflow execution. (A) Example prompt requesting installation of the asari framework, automatic MCP server generation, and exposure of its capabilities through ReCo, together with the corresponding ReCo response summarizing the generated tools and usage instructions. (B) Representative prompt requesting LC-MS metabolomics processing of the SZ22 dataset and empirical-compound annotation, with the corresponding ReCo response reporting processing statistics and quality metrics. (C) Automatically generated output artifacts, including processing results and quality-control summary files produced by the asari MCP server. (D) Representative workflow outputs comprising the detected composite feature map and an extracted-ion chromatogram linked to a representative mass track. (E) Feature-quality overview showing the distribution of detected LC-MS features according to goodness of peak shape, peak area, and chromatographic selectivity. (F) Distributions of key quality-control metrics (signal-to-noise ratio, goodness-of-fit, and chromatographic selectivity) computed from the processed dataset.

### Monte Carlo Radiation Dosimetry Framework Ingestion and Functional Testing

For DosimeTron, ReCo ingested a local multi-server agentic dosimetry codebase and preserved its original functional separation by generating four MCP servers corresponding to DICOM handling, preprocessing, GATE MC execution, and organ/tissue-specific radiation dose reporting. The resulting ReCo installation exposed 24 tools, including one additional job-listing tool, and the servers passed live MCP initialization and tool-list checks. Functional evaluation used six PET/CT cases, including both 18F and 68Ga radiotracers, and executed the complete scan-to-dose-report workflow through the generated DosimeTron MCP servers. ReCo automatically detected the radionuclide, prepared the required inputs, executed the MC workflow, and generated organ-level dosimetry reports and segmentation-derived organ statistics. Quantitative comparison against the original DosimeTron agentic runs showed near-identical re-hosting of the pipeline across 651 organ observations (Figure 4), with Pearson correlation and Lin concordance correlation both equal to 0.99999 and mean absolute percentage difference of 0.37% for scan-time organ dose rate. Input cumulated activity values matched to six significant figures, and segmentation-derived organ volume, mass, and voxel-count values were reproduced with negligible differences. These results demonstrate that ReCo could re-host the original DosimeTron workflow with high numerical fidelity under the tested conditions.

**Figure 4.**
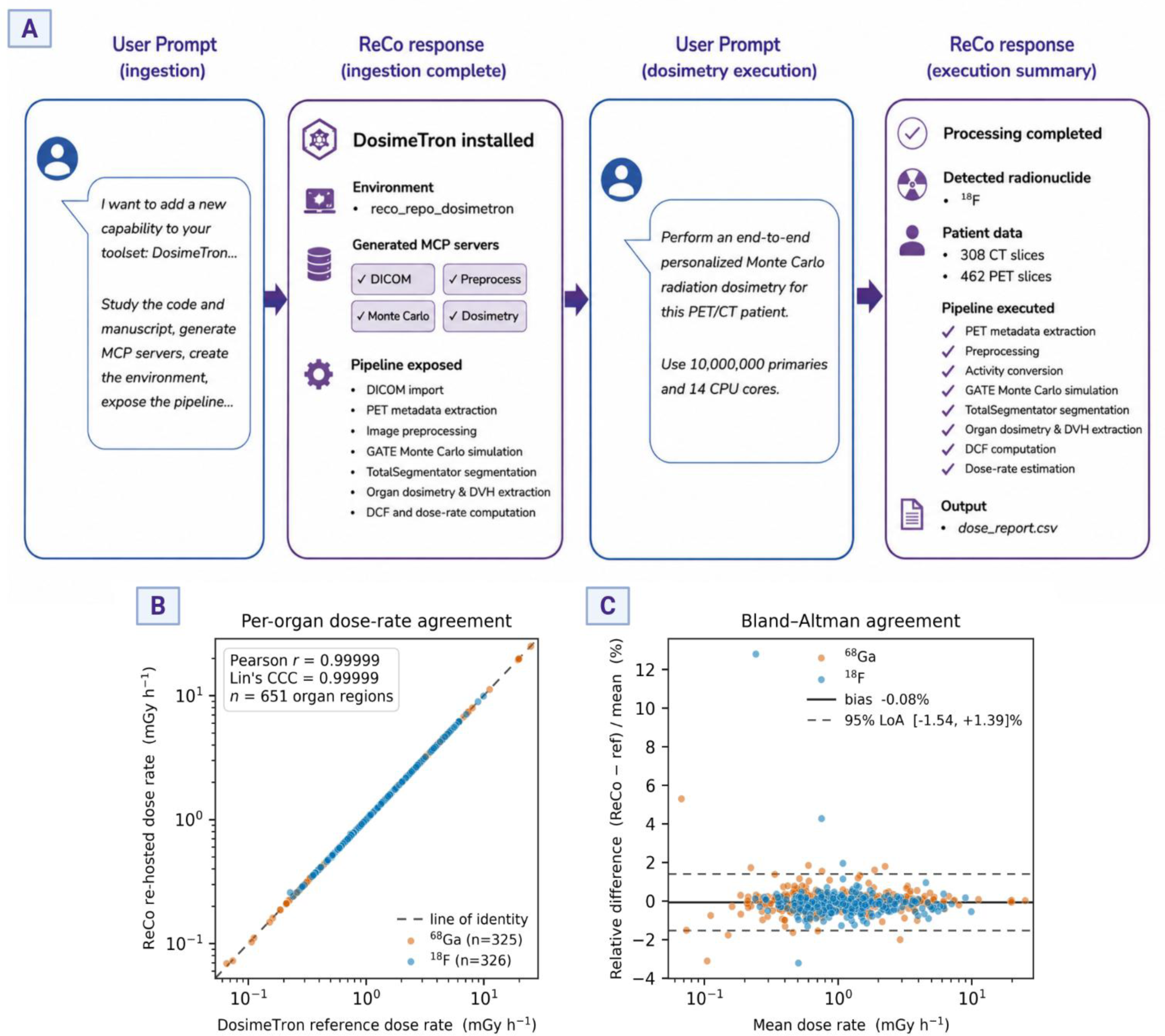
(A) ReCo automatically ingests the DosimeTron PET/CT MC dosimetry framework, generates MCP servers exposing the complete dosimetry pipeline, and subsequently executes an end-to-end personalized dosimetry request. (B) Agreement between organ dose rates generated through the ReCo-executed workflow and the reference DosimeTron pipeline across all evaluated organs. (C) Bland– Altman analysis demonstrating negligible bias and narrow limits of agreement, confirming faithful execution of the original MC dosimetry workflow through the automatically generated MCP interface.

### Orthanc Infrastructure Setup and Orchestration

For Orthanc, ReCo configured a local DICOM server from scratch using Docker, enabled REST and DICOMweb access, diagnosed and corrected an initial plugin-configuration issue, and generated an MCP client with tools for server inspection, resource search, metadata retrieval, DICOM download, DICOM upload, and job monitoring. ReCo then used Orthanc as a PACS-like coordination layer. In the setup experiment, a PET/CT study was uploaded to Orthanc, retrieved through the generated MCP client, converted for downstream processing, segmented, and returned to Orthanc as DICOM RTSTRUCT objects. In the orchestration experiments, ReCo used Orthanc-mediated data access to run DosimeTron workflows, Merlin workflows, and TotalSegmentator workflows, then pushed the resulting artifacts back to Orthanc as DICOM RTSTRUCT, RTDOSE, Structured Report, and document objects (Figure 5). The live Orthanc inventory confirmed the presence of the pushed-back object classes, and generated DICOM files were checked for validity. For the DosimeTron-based workflow, comparison with the direct file-based validation runs showed that PACS-mediated data access did not introduce dosimetric drift in the tested studies. The Orthanc evaluation therefore demonstrated that ReCo’s self-extension mechanism could be applied not only to scientific model wrappers, but also to infrastructure that enables multi-framework AI workflows in a clinically realistic imaging-data environment.

**Figure 5.**
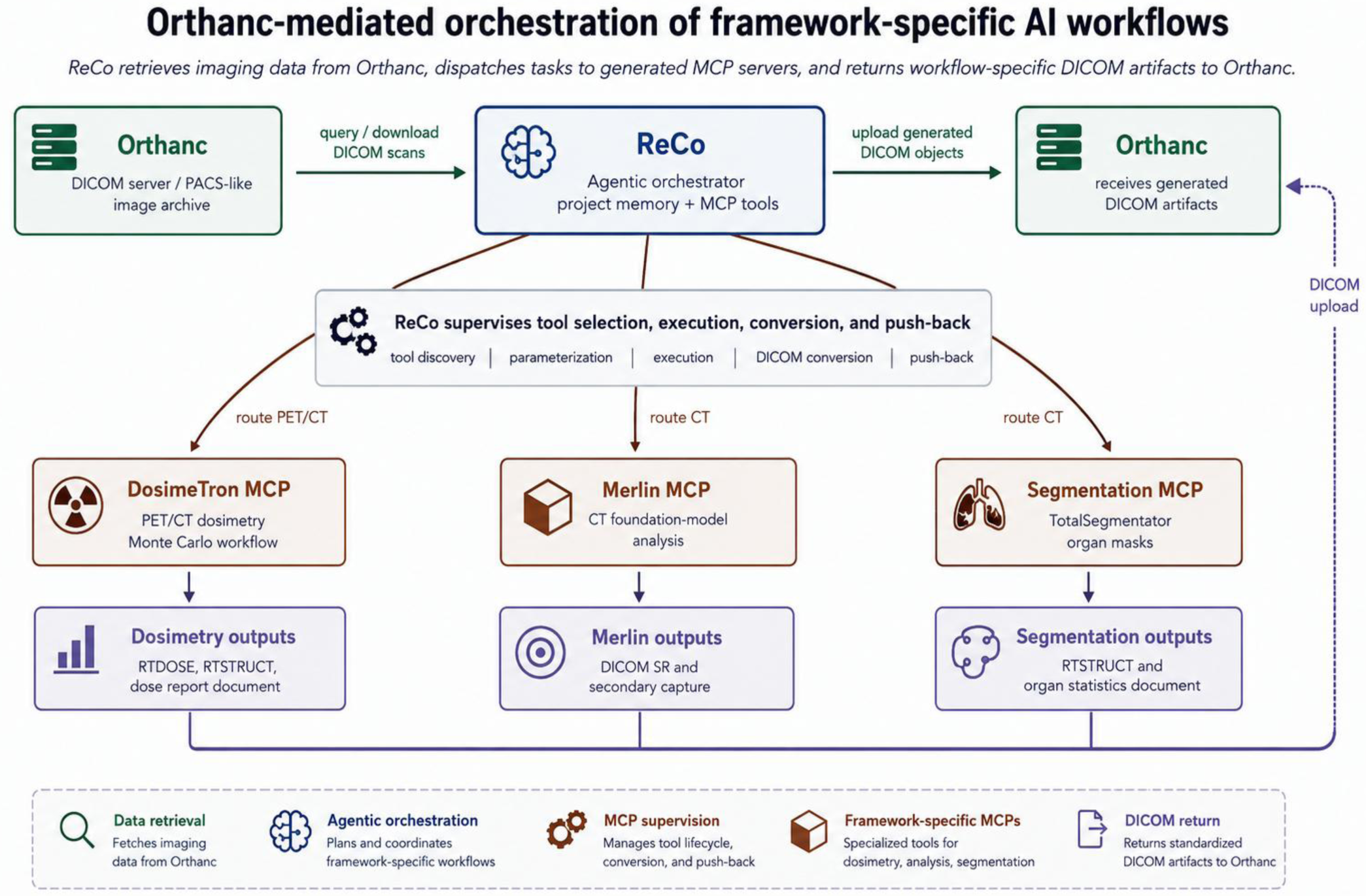
ReCo-orchestrated workflow in which imaging data are retrieved from Orthanc, routed through framework-specific MCP servers for DosimeTron radiation dosimetry, Merlin CT foundation-model analysis, and segmentation, and uploaded back to Orthanc as DICOM artifacts.

## Discussion

ReCo was developed as a research-oriented agentic platform that combines a preconfigured toolkit with a mechanism for expansion beyond its original capabilities. The system bundles MCP servers for established research tasks [7, 8], but its main value is not limited to this predefined toolset. ReCo was designed to (i) study new frameworks from repositories, manuscripts, or local codebases, (ii) identify their dependencies and execution patterns, (iii) create isolated runtime environments, (iv) expose their functions through MCP servers, (v) verify the generated interfaces, and (vi) produce documentation that helps users understand the newly added capabilities. This design addresses a practical limitation in contemporary computational/AI research, that new tools are released rapidly, but their adoption often requires programming expertise, dependency management, environment troubleshooting, and detailed knowledge of each framework’s interface. By automating this configuration and encapsulation process, ReCo can reduce the technical burden associated with adopting new research software and can help researchers keep pace with an expanding ecosystem of AI and computational frameworks. This may be particularly valuable for researchers with limited programming experience, who can interact with advanced tools through a natural-language interface without manually resolving dependency conflicts or writing framework-specific wrappers. In addition, ReCo has potential educational value as a training platform for medical doctors, medical physicists, biomedical engineers, and other researchers learning AI development, deployment, evaluation, and implementation, because each ingested framework is not only made executable but also documented and presented as part of an inspectable research workflow.

ReCo’s self-extension capability was not limited to wrapping AI models or command-line research frameworks. The Orthanc evaluation demonstrated that the same mechanism can be applied to imaging infrastructure, allowing ReCo to configure a PACS system, generate a dedicated MCP client for communication with the DICOM server, retrieve imaging examinations, execute AI or computational workflows, and return generated artifacts to the imaging archive. This addresses an important translational barrier, that many AI models remain confined to research environments not only because of model-performance limitations, but also because integration with clinical imaging infrastructure requires DICOM handling, PACS communication, data conversion, output packaging, and workflow coordination. In the evaluated setting, ReCo used Orthanc as a PACS-like environment and combined it with newly ingested or bundled capabilities, including DosimeTron workflows, Merlin CT foundation-model analysis, and segmentation workflows, before returning workflow-specific DICOM objects to the server. This suggests that ReCo can function as an integration layer between research software and institutional imaging ecosystems, while preserving user supervision and auditability. The same property is relevant for in-house tools developed by clinical scientists or research groups. ReCo can ingest such local frameworks, document their capabilities, expose them through MCP servers, and make them interoperable with other tools in the ReCo ecosystem. In this way, locally developed methods can be combined with external foundation models, segmentation models, dosimetry pipelines, or DICOM infrastructure, supporting more complex research workflows and providing a path toward testing how these tools could operate within the broader environment that handles patient data.

The study has certain limitations. First, ReCo was evaluated on a limited number of frameworks, selected to span different software archetypes and research domains but still representing only a small sample of the broader research-software ecosystem. Future work should evaluate the system across a larger set of frameworks, domains, and research groups, including externally defined real-world use cases in which teams attempt to ingest and use tools relevant to their own scientific workflows. Second, all experiments were performed on a single workstation. Although ReCo adapted to the constraints of this environment, including GPU-memory limitations and framework-specific runtime requirements, additional testing is needed to characterize its behavior across different hardware configurations, accelerator types, and institutional computing environments. Third, although ReCo supports selection among different hosted and local reasoning backends, all validation experiments in this study used Claude Opus 4.8 as the core reasoning model. The extent to which the same ingestion, planning, troubleshooting, documentation, and orchestration behavior is preserved with other large language models remains to be evaluated. This is particularly relevant for smaller local models, because successful operation with local reasoning backends would support more private and fully local deployments.

## Conclusion

This study presents ReCo, an agentic research platform designed for self-configuration and self-extension. By combining a bundled MCP toolkit, structured skills, project memory, and a desktop user interface, ReCo provides a unified environment for configuring, documenting, and operationalizing complex computational and AI frameworks. The validation experiments showed that ReCo can ingest heterogeneous research software, expose newly added capabilities through MCP servers, and connect these capabilities to imaging infrastructure in clinically realistic workflows. In doing so, ReCo can reduce technical barriers for researchers with limited programming experience and support faster adoption of emerging research tools.

## Code Availability

The official open-source repository for the ReCo framework is available at: https://github.com/eltzanis/ReCo

## Competing Interests

The authors declare no competing interests.

## Data Availability

All data produced in the present study are available upon reasonable request to the authors.

https://github.com/eltzanis/ReCo

## Acknowledgments

None.

## Funding

None.

## Notes

### Competing Interest Statement

The authors have declared no competing interest.

## References

[1] Yao S, Zhao J, Yu D, et al. ReAct: Synergizing reasoning and acting in language models. arXiv [cs.CL] 2022. https://arxiv.org/abs/2210.03629

[2] Schick T, Dwivedi-Yu J, Dessì R, et al. Toolformer: Language models can teach themselves to use tools. arXiv [cs.CL] 2023. https://arxiv.org/abs/2302.04761

[3] Ferber, D., Hilgers, L., Höper, C. et al. Towards autonomous medical artificial intelligence agents. Nature (2026). 10.1038/s41586-026-10675-5

[4] Tzanis E, Adams LC, Akinci D’Antonoli T, et al. Agentic systems in radiology: Principles, opportunities, privacy risks, regulation, and sustainability concerns. Diagn Interv Imaging. 2026;107(1):7–16. 10.1016/j.diii.2025.10.002

[5] Liévin, V., Palepu, A., Weng, WH. et al. Towards Conversational AI for Disease Management. Nature (2026). 10.1038/s41586-026-10764-5

[6] Introducing the Model Context Protocol. https://www.anthropic.com/news/model-context-protocol. 2024. Accessed 11/7/2026.

[7] Tzanis E, Klontzas ME. ReclAIm: A multiagent framework for monitoring and correcting performance decline in medical imaging AI. Radiology: Artificial Intelligence. 2026;8(4):e250923. 10.1148/ryai.250923

[8] Tzanis E, Klontzas ME. mAIstro: An open-source multi-agent system for automated end-to-end development of radiomics and deep learning models for medical imaging. European Journal of Radiology Artificial Intelligence. 2025;4:100044. 10.1016/j.ejrai.2025.100044

[9] Claude Agent SDK. https://code.claude.com/docs/en/agent-sdk/overview. Accessed 11/7/2026.

[10] Wasserthal J, Breit H-C, Meyer MT, et al. TotalSegmentator: Robust Segmentation of 104 Anatomic Structures in CT Images. Radiology: Artificial Intelligence. 2023;5(5). 10.1148/ryai.230024

[11] I Isensee, F., Jaeger, P.F., Kohl, S.A.A., et al. nnU-Net: a self-configuring method for deep learning-based biomedical image segmentation. Nat Methods 18, 203–211 (2021). 10.1038/s41592-020-01008-z

[12] Anaconda.org. https://anaconda.org/channels/anaconda/packages/conda/overview. Accessed 11/7/2026.

[13] Anthropic. https://www.anthropic.com/. Accessed 11/7/2026.

[14] Deepseek. https://www.deepseek.com/en/. Accessed 11/7/2026.

[15] Ollama. https://ollama.com/. Accessed 11/7/2026.

[16] Electron. https://www.electronjs.org. Accessed 11/7/2026.

[17] React. https://react.dev. Accessed 11/7/2026.

[18] Blankemeier, L., Kumar, A., Cohen, J.P., et al. Merlin: a computed tomography vision– language foundation model and dataset. Nature 652, 1318–1328 (2026). 10.1038/s41586-026-10181-8

[19] Zhao C, Guo P, Yang D, et al. MAISI-v2: Accelerated 3D high-resolution medical image synthesis with rectified flow and region-specific contrastive loss. arXiv [cs.CV] 2025. 10.48550/arXiv.2508.05772

[20] Li, S., Siddiqa, A., Thapa, M. et al. Trackable and scalable LC-MS metabolomics data processing using asari. Nat Commun 14, 4113 (2023). 10.1038/s41467-023-39889-1

[21] Tzanis E, Klontzas ME, Tzortzakakis A. DosimeTron: Automating personalized Monte Carlo radiation dosimetry in PET/CT with agentic AI. arXiv [physics.med-ph] 2026. 10.48550/arXiv.2604.06280

[22] Orthanc. https://www.orthanc-server.com/. Accessed 11/7/2026.

[23] Wasserthal, J. & Akinci D’Antonoli, T. TotalSegmentator MRI dataset: 298 MRI images with segmentations for 56 anatomical regions (Version 1.0.0) [Dataset]. Zenodo 2024. 10.5281/zenodo.11367005

[24] Jeblick, K., Schachtner, B., Mittermeier, A., et al. A whole-body PSMA-PET/CT dataset with manually annotated tumor lesions (Version 2) [dataset]. The Cancer Imaging Archive 2026. https://www.cancerimagingarchive.net/collection/PSMA-PET-CT-Lesions/. Accessed 11/7/2026.

